# Using Bayesian latent class models to evaluate tuberculosis diagnostics in a pediatric cohort study in Kenya

**DOI:** 10.64898/2026.11.17.26368380

**Authors:** Jerphason Mecha, George Muhua, Joy Githua, Jaclyn N. Escudero, Lucy Kijaro, Lilian N. Njagi, Grace John-Stewart, Elizabeth Maleche-Obimbo, Videlis Nduba, Sylvia M. LaCourse, Lurdes Y.T. Inoue

**Author notes:** **Corresponding author**: Jerphason Mecha, BOX 54840-00200 Nairobi-Kenya. Equal contribution: LYT Inoue, SM LaCourse.

## Abstract

**Background:** Pediatric tuberculosis (TB) remains difficult to diagnose. Culture, the gold standard, has limited sensitivity in children (∼40-60%). We analyzed 452 symptomatic children ≤15 years who underwent chest x-ray (CXR), respiratory sample testing with Xpert Ultra, smear microscopy and culture, tuberculin skin test (TST) and interferon gamma release assay (IGRA) in Nairobi, Kenya.

**Methods:** Three Bayesian latent class models (BLCMs) were used: Model 1 assumed conditional test independence; Model 2 incorporated random effects to account for shared biological (Smear, Xpert Ultra and culture) or immunologic (IGRA, TST) test dependence; Model 3 extended Model 2 by adjusting for clinical covariates and NIH TB signs and symptoms. We estimated TB prevalence, test sensitivities/specificities and covariate effects.

**Results:** Prevalence estimates under Models 1 [20.2% (15.9–24.7)] and 2 [24.5% (19.6–30.0)] were higher than under Model 3 [14.8% (11.0–19.3)]. All the three model-based prevalence estimates were considerably higher than the 9.9% of children identified through microbiological testing alone. Sensitivity and specificity were broadly consistent across models. Model 3 was considered preferred because it accounted for dependence among the tests and adjusted prevalence and sensitivity for clinical covariates. Under this Model 3, culture and GeneXpert MTB/RIF Ultra demonstrated moderate sensitivity (56.9% and 63.2%, respectively), while smear microscopy showed lower sensitivity (29.8%). CXR achieved moderate sensitivity (79.7%) but limited specificity (40.2%). IGRA and TST displayed sensitivities of 63.7% and 50.1%, and specificities of 81.5% and 65.8%. Older age was positively associated with TB (log-odds ratio 1.09; 95% CrI: 0.14–2.18).

**Conclusion:** BLCMs estimated TB prevalence and test accuracies without a gold standard. Sensitivity analyses confirmed robustness to priors. Radiography demonstrated moderate sensitivity while culture and GeneXpert retained excellent specificity with improved test performance with older age.

## Introduction

Tuberculosis (TB) remains a leading global cause of morbidity and mortality(1). Children account for 10–15% of TB cases in high-burden settings(2,3). Malnutrition and HIV infection further increase susceptibility to TB and complicate diagnosis and management(4).

Accurate diagnosis of TB in children remains a major challenge due to non-specific symptoms, difficulties obtaining sputum, and the paucibacillary nature of pediatric disease(5,6). Culture, the best available reference standard, achieves only 40–60% sensitivity (5). Molecular diagnostics such as geneXpert have improved time to TB detection, yet their accuracy is usually evaluated against culture, an imperfect gold standard(7).

Consensus clinical case definitions attempt to standardize pediatric TB diagnosis by integrating clinical, radiologic, microbiologic, and TB exposure data(8), but rely heavily on subjective interpretation of clinical and radiographic findings and may increase variability(9). Given limitations of both microbiological tests and composite clinical algorithms, there is an urgent need for analytic methods that more accurately estimate pediatric TB prevalence and test performance.

By modeling true TB status as an unobserved latent variable and jointly analyzing results from multiple tests, Bayesian Latent Class Models (BLCM) allow estimation of test sensitivity, specificity, and disease prevalence (10,11). Application of BLCM in pediatric TB remains limited (12,13). Existing studies reviewed a narrower panel of diagnostic tests, did not incorporate immunologic assays (i.e. QFT), and insufficiently adjust for covariates that may influence test performance.

We applied BLCM to a comprehensive set of diagnostic tests including microbiologic, immunologic, radiologic and NIH defined TB signs and symptoms for pediatric TB. We incorporated key covariates and evaluated sensitivity to prior specifications.

## Methods

### Study Design and Population

We analyzed cross-sectional data collected at enrollment from May 5, 2022 to April 8, 2025 in a prospective cohort of a pediatric TB diagnostic study in Nairobi, Kenya(14). Children under 15 years of age presenting with symptoms underwent microbiological, immunological, and radiological evaluation for suspected pulmonary TB. Both HIV-positive and HIV-negative children were included. We excluded children who had received >7 days of TB treatment prior to enrollment.

### Study Procedures

Participants underwent standardized clinical evaluation with structured interviews and physical examination. Positive TST was defined as induration ≥5mm in children with HIV or malnourished and ≥10 mm otherwise. Respiratory samples were collected for geneXpert, smear microscopy, and mycobacterial culture using the Mycobacteria Growth Indicator Tube (MGIT). CXRs were independently reviewed by a radiologist and study clinician; discrepancies were resolved by a pediatric pulmonologist. CXR findings were classified as suggestive or non-suggestive of TB, and abnormalities categorized according to standardized criteria(15). Blood was collected for the QuantiFERON-TB Gold Plus (QFT-Plus) assay to assess *M. tuberculosis* infection, with results interpreted per manufacturer’s instructions. Nutritional status was assessed using weight-for-age z-scores (WAZ) per WHO growth standards. Participants were classified into NIH-defined categories: Confirmed TB (microbiological confirmation), Unconfirmed TB (symptoms plus supportive evidence but negative microbiological results), and Unlikely TB (15).

### Heuristic TB Model

We developed a heuristic TB model to represent TB pathophysiology assumptions and biological mechanisms underlying diagnostic outcomes. This model extends the framework developed by Schumacher et al. (2016) by including immunologic tests for TB infection (QFT) and covariates such as MUAC and WAZ. Microbiological tests were assumed to reflect bacillary load, creating potential statistical dependence among tests. Immunological tests (TST and QFT) were modeled as influenced by host immune status and disease severity, with anticipated reductions in sensitivity among HIV-infected or malnourished children. CXR was assumed to have moderate specificity due to overlap with other pulmonary diseases. Covariates including HIV status, age, nutritional indicators (MUAC, WAZ, BMI), and household TB exposure were expected to influence test performance and TB prevalence **Figure 1**.

**Figure 1:**
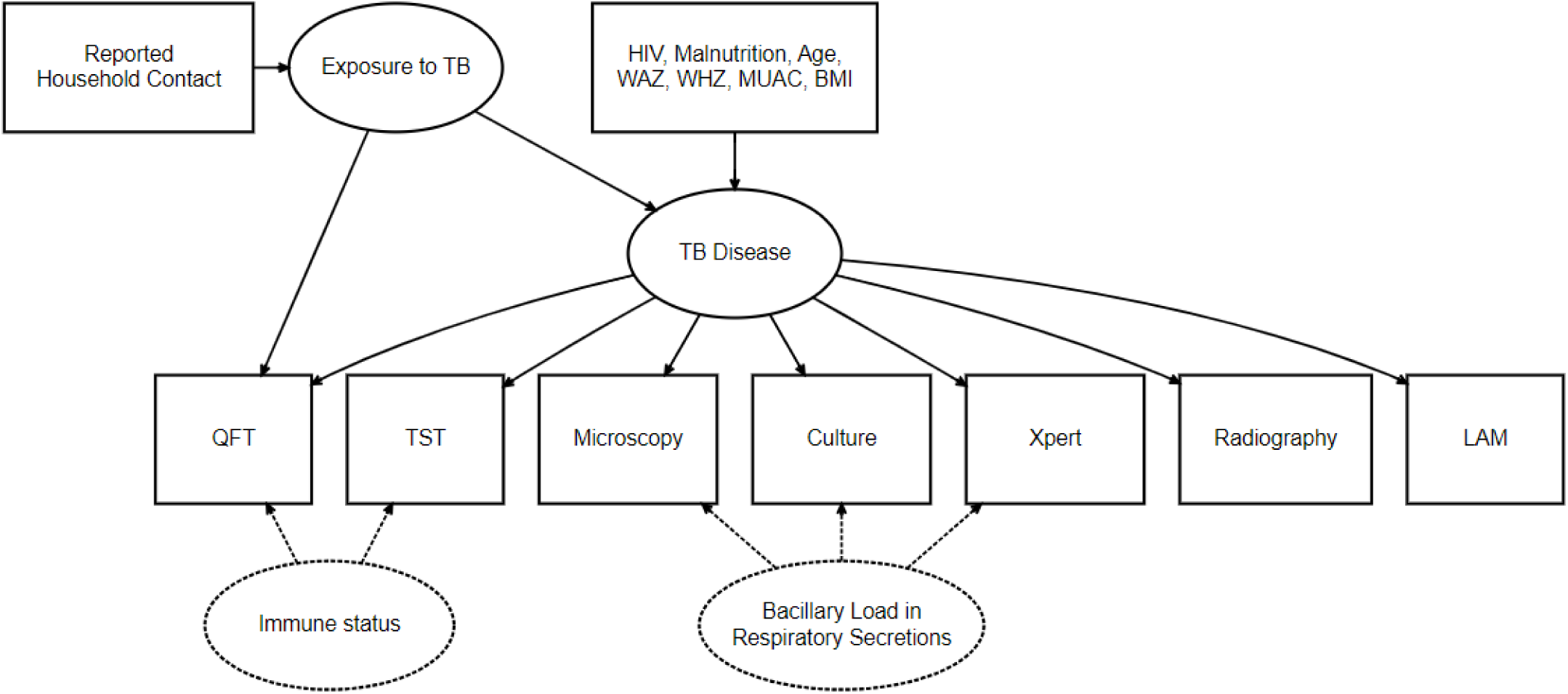
Heuristic model specifying prior beliefs of relationships between latent and observed variables. Nodes for immune status and bacillary load are random effects.

### Bayesian Estimation and modeling of latent class models

We used Bayesian methods to estimate latent class model parameters, incorporating prior knowledge on diagnostic test performance and disease prevalence. The likelihood function was constructed based on assumptions about the different tests. We developed 3 different models; Model 1 assumed that, given the true TB status, the diagnostic tests were conditionally independent. In Model 2, we introduced a continuous random effect to represent bacillary load in respiratory secretions. This random effect accounted for the shared dependence between microbiological tests and allowed the sensitivities of these tests to vary depending on the bacillary load. Model 3 builds on Model 2 by adding covariates affecting both TB prevalence and test sensitivity. We used logistic regression to model the probability of TB as a function of covariates. In addition, we explicitly modeled the link between bacillary load and TST sensitivity using a quadratic term to allow for nonlinear effects. Each test outcome was then modeled with a Bernoulli distribution, conditional on the child’s true TB status. Description of the 3 models with respective model strings and prior specification are outlined in the **supplemental materials 1,2 respectfully**.

## RESULTS

### Baseline Characteristics

Among the 452 children enrolled in the pediatric TB diagnostic study, 238 (52.7%) were female, median age was 2.5 years (IQR: 1.1–5.3). The prevalence of HIV-exposed uninfected children was 20 (4.4%), while 21 (4.6%) were children living with HIV(CLHIV). The median WAZ was –1.6 (IQR: –2.8, –0.5). Most participants 369 (81.6%) met the NIH-defined criteria for TB signs and symptoms, and over one-third (37.6%) reported a household contact with TB. Regarding diagnostic testing, 18 (4.6%) were culture positive, 39 (9.2%) tested positive on GeneXpert MTB Ultra/RIF, and 14 (3.4%) on smear microscopy. Overall, 9.9% of the children had microbiological confirmation of TB(Positive culture or Positive GeneXpert MTB Ultra/RIF). Radiographic abnormalities suggestive of TB were observed in 281 (62.2%). Immunologic testing yielded positive results in 75 (21.9%) for QuantiFERON-TB Gold Plus and 153 (34.5%) for tuberculin skin test. Urine LAM was positive in 29 (12.9%) of children (**Table 1** shows demographic, clinical diagnostic characteristics of the pediatric TB cohort).

**Table 1:**
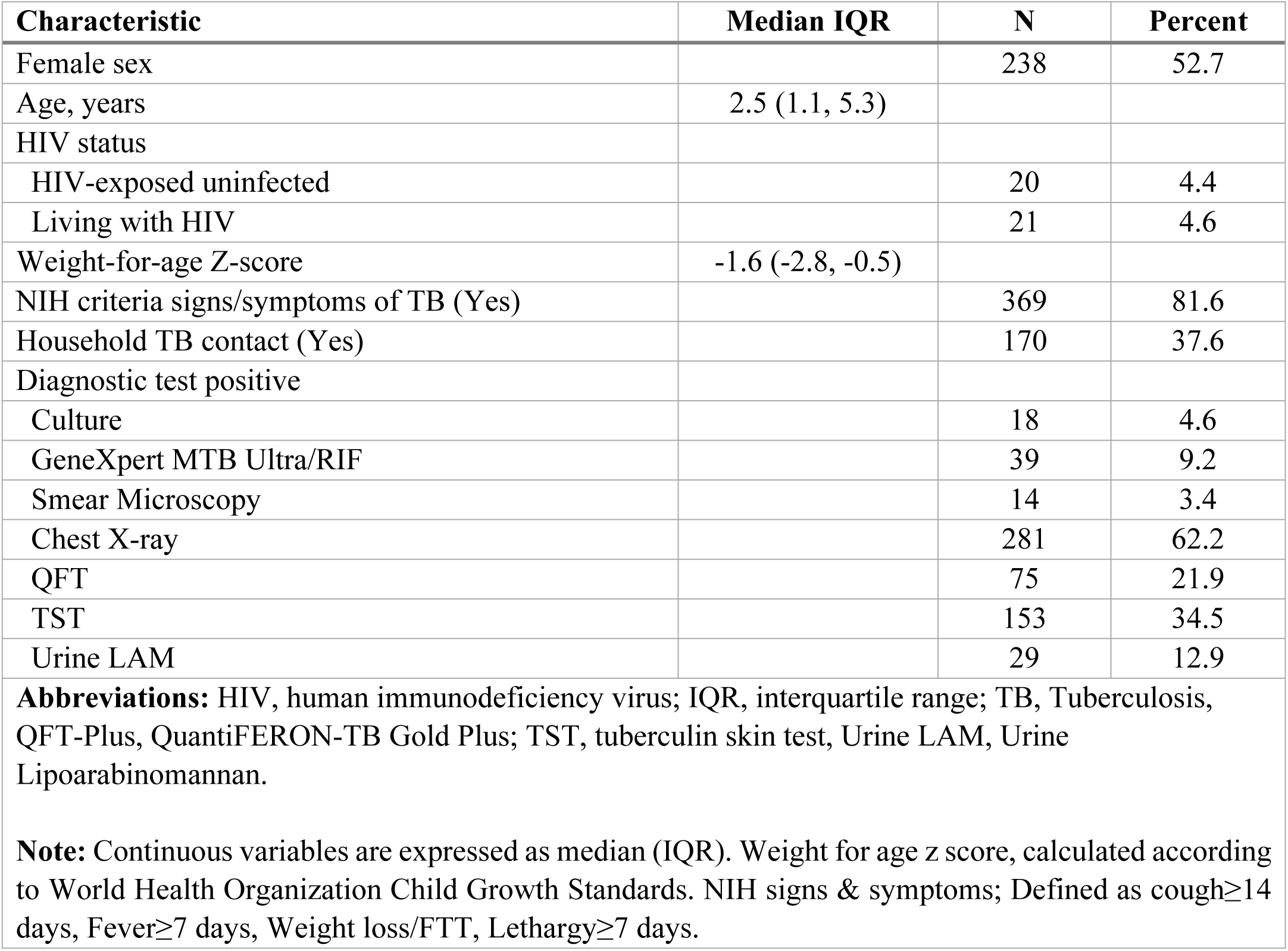
Demographic, Clinical and Diagnostic characteristics of Children Enrolled in the TB Diagnostic Cohort (N=452)

### TB Prevalence Estimates

Under Model 1, the posterior median TB prevalence was 20.2% (95% CrI: 15.9–24.7%). The inclusion of shared dependence structures in Model 2 produced a slightly higher estimate of 24.5% (95% CrI: 19.6–30.0%) (**Table 2**).

**Table 2:** Posterior estimates (posterior mean and limits of the 95% posterior credible interval) of the prevalence, test sensitivity and specificity parameters under models 1,2 and 3.

| **Test** | **Parameter** | **Model 1** | **Model 2** | **Model 3** |
|---|---|---|---|---|
|  | Prevalence | 20.2 (15.9, 24.7) | 24.5 (19.6, 30.0) | 14.8 (11.0, 19.3) |
| **Culture** | Sensitivity | 52.1 (40.8, 64.3) | 51.6 (39.7, 63.5) | 56.9 (46.0, 67.1) |
|  | Specificity | 99.2 (98.0, 99.9) | 99.1 (97.7, 99.9) | 98.0 (96.3, 99.2) |
| **Xpert** | Sensitivity | 63.1 (52.1, 73.4) | 60.0 (48.6, 71.0) | 63.2 (53.3, 72.4) |
|  | Specificity | 98.0 (95.6, 99.7) | 96.9 (94.2, 99.0) | 95.2 (92.6, 97.3) |
| **Microscopy** | Sensitivity | 31.4 (22.3, 41.5) | 26.6 (17.5, 37.3) | 29.8 (21.1, 39.7) |
|  | Specificity | 99.2 (97.9, 99.9) | 99.0 (97.6, 99.8) | 98.1 (96.5, 99.2) |
| **Chest X-ray** | Sensitivity | 82.9 (68.6, 94.0) | 77.4 (61.6, 91.4) | 79.7 (59.5, 93.5) |
|  | Specificity | 40.5 (35.6, 45.5) | 40.7 (35.1, 46.3) | 40.2 (34.8, 45.6) |
| **QFT** | Sensitivity | 57.3 (38.6, 76.4) | 63.3 (43.4, 81.9) | 63.7 (29.8, 90.5) |
|  | Specificity | 83.0 (78.3, 87.6) | 87.0 (81.3, 92.3) | 81.5 (76.2, 87.2) |
| **TST** | Sensitivity | 57.2 (40.5, 74.7) | 64.1 (46.5, 80.6) | 50.1 (20.0, 82.6) |
|  | Specificity | 67.9 (62.9, 72.7) | 70.9 (65.2, 76.5) | 65.8 (60.2, 72.4) |
| **Model Fit** | DIC | 1906.2 | 2014.9 | 4251.2 |

**Abbreviations**; QFT-QuantiFERON-TB Gold Test , TST-Tuberculin Skin Test, DIC- Deviance Information Criteria, Xpert-Xpert MTB/ULTRA

### Diagnostic Test Performance—Model 1 (Conditional Independence)

Model 1 yielded moderate-to-high sensitivity estimates for molecular and culture-based diagnostics. Xpert demonstrated a sensitivity of 63.1% (95% CrI: 52.1–73.4%), while culture showed a sensitivity of 52.1% (95% CrI: 40.8–64.3%). Microscopy had lower sensitivity at 31.4% (95% CrI: 22.3–41.5%). Immunologic assays performed moderately: TST sensitivity was 57.2% (95% CrI: 40.5–74.7%) with a specificity of 67.9% (95% CrI: 62.9–72.7%), while QFT showed similar sensitivity at 57.3% (95% CrI: 38.6–76.4%) but higher specificity at 83.0% (95% CrI: 78.3–87.6%). CXR retained high sensitivity at 82.9% (95% CrI: 68.6–94.0%) but low specificity at 40.5% (95% CrI: 35.6–45.5%) (**Table 2**).

### Diagnostic Test Performance Model 2 (Conditional dependence)

In Model 2, the addition of latent random effects to capture shared biological and immunologic dependence resulted in decreased sensitivity estimates for microbiological tests. Baseline culture sensitivity declined to 51.6% (95% CrI: 39.7–63.5%), Xpert to 60.0% (95% CrI: 48.6–71.0%), and microscopy to 26.6% (95% CrI: 17.5–37.3%).

Baseline sensitivity and specificity of immunologic and radiographic tests remained similar to Model 1 because their likelihood structures were not functions of the random effects. CXR remained similarly sensitive 77.4% (95% CrI: 61.6–91.4%), with poor specificity 40.7% (95% CrI: 35.1–46.3%), while TST and QFT sensitivities remained stable (64.1% (95% CrI: 46.5–80.6%) and 63.3% (95% CrI: 43.4–81.9%), respectively). Full posterior estimates for Models 1 and 2 are shown in **Table 2**.

### Influence of Covariates on TB Prevalence and Diagnostic Test Performance (Model 3) TB Prevalence Estimates

Model 3 incorporated multiple covariates, including HIV status, malnutrition indicators (WAZ, MUAC), household TB exposure, and clinical symptoms. Covariate adjustment produced a lower TB prevalence (at the average values of the covariates) of 14.8% (95% CrI: 11.0–19.3%), representing a 26.7% reduction from Model 1 and a substantially narrower credible interval.

### Diagnostic Test Performance—Model 3

Microbiological test performance was relatively stable or showed modest increases after covariate adjustment. Sensitivities were 56.9% (95% CrI: 46.0–67.1%) for culture, 63.2% (95% CrI: 53.3–72.4%) for Xpert, and 29.8% (95% CrI: 21.1–39.7%) for microscopy, with all maintaining specificities >95%. The positive shared random effect coefficient (estimate 3.01; 95% CrI: –0.66 to 4.92) quantified improved microbiologic detection with increasing bacillary burden.

Immunologic tests showed variable performance. TST sensitivity declined to 50.1% (95% CrI: 20.0–82.6%), while QFT increased to 63.7% (95% CrI: 29.8–90.5%), both with wide credible intervals driven by uncertainty in covariate effects such as malnutrition (QFT: estimate 0.35 (95% CrI: -2.31–3.32%); TST: estimate 0.20 (95% CrI: -2.06–2.77%) and HIV status (QFT: estimate 0.01 (95% CrI: -6.14–6.20%); TST: estimate –0.44 (95% CrI: -6.41–5.81%). A quadratic random-effect term (estimate 1.58; 95% CrI: 0.06–4.30) suggested potential nonlinear associations between bacillary load and immune assay positivity. CXR remained moderately sensitive 79.7% (95% CrI: 59.5–93.5%), with low specificity 40.2% (95% CrI: 34.8–45.6%). The influence of covariates across tests is illustrated in **Table 2**; **Figure 2**.

**Figure 2:**
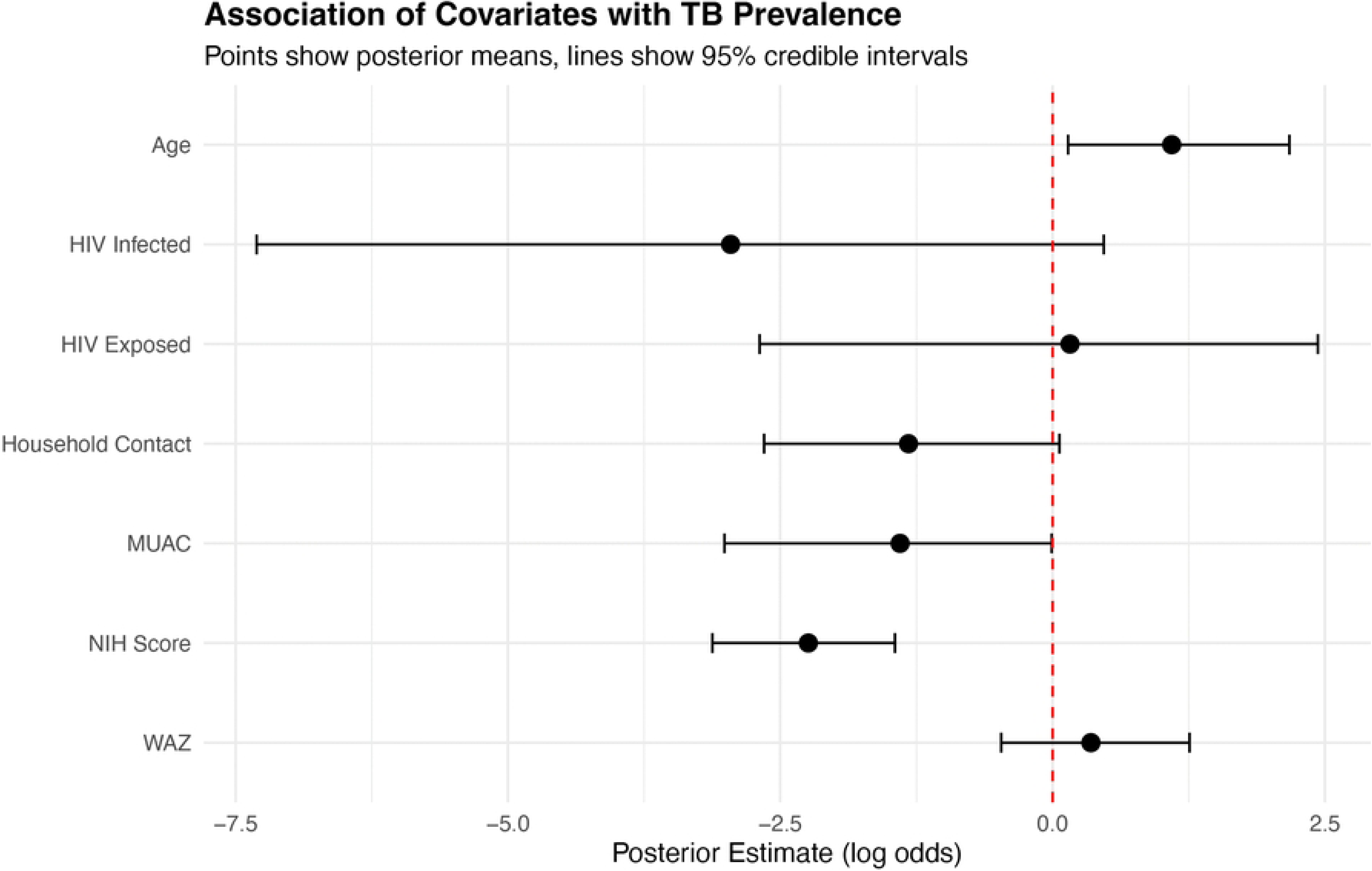
Association of covariates with TB prevalence.

Convergence diagnostics indicated satisfactory performance for all three models, with R-hat values close to 1.00, Geweke Z-scores centered near zero and well mixed trace plots **(Supplemental material 3, Figures 1-3**, **Supplemental material 3, Table 1**). Deviance Information Criteria under informative priors favored model 1(1906.2) over model 2 (2014.9), while model 3 showed substantially higher DIC (4251.2), reflecting its greater complexity and the sparseness of covariate data rather than inferior clinical utility (**Supplemental material 4, figure 1**).

**Figure 3:**
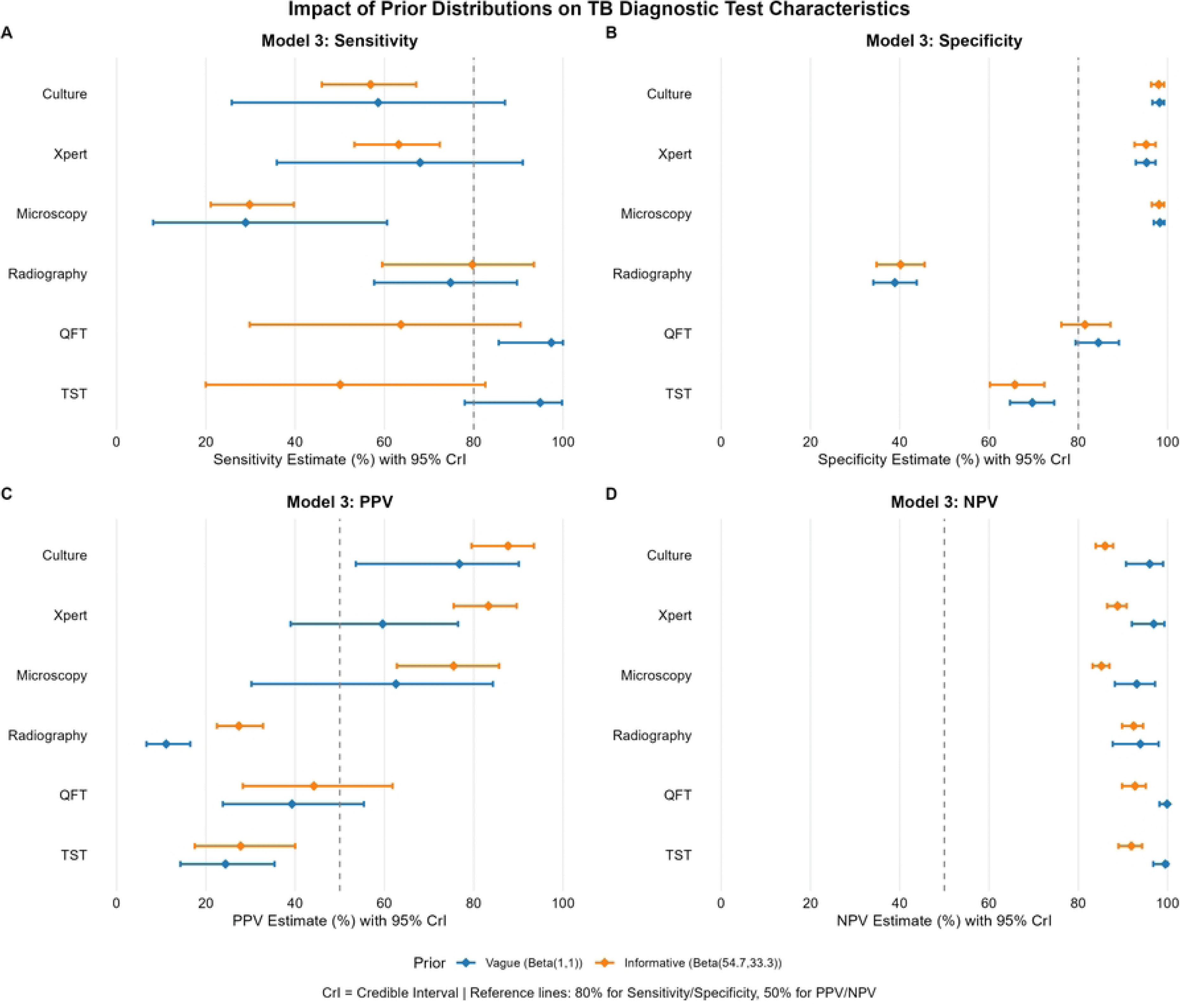
Imapact of prior specification on diagnostictest performance.

### Impact of Prior Distributions on Diagnostic Test Characteristics Test Sensitivity

Informative priors reduced uncertainty for microbiological tests, especially culture, whose sensitivity estimate (∼57%) remained stable with a narrower CrI. geneXpert displayed similar improvements in precision of its sensitivity estimate. Microscopy’s sensitivity was robust to prior choice, with point estimates remaining at ∼29% but narrower intervals under informative priors. In contrast, QFT and TST modeled with vague priors showed marked shifts when other model components adopted informative priors. QFT sensitivity decreased from 97.4% to 63.7%, and TST from 94.9% to 50.1%, demonstrating strong dependence on prior prevalence. CXR estimates were unaffected.

### Test Specificity

Specificity estimates were more stable. Culture specificity remained >98% regardless of prior choice, with informative priors slightly improving precision. Xpert and microscopy also had narrower CrIs under informative priors. QFT and TST had small downward shifts (QFT: 84.5% lowered to 81.5%; TST: 69.7% lowered to 65.8%), reflecting system-wide prior influences rather than test-specific data (**Figure 3**).

### Predictive Values

Predictive values were strongly sensitive to prior specifications. PPV for culture, Xpert, and microscopy increased substantially (e.g., culture: 76.8% increased to 87.7%; Xpert: 59.6% increased to 83.3. CXR PPV more than doubled (11.1% increased to 27.4%). QFT and TST similarly showed increased PPV. Negative predictive values (NPV) decreased across all tests, most notably for QFT and TST (both >99% lowered to ∼92%), underscoring the sensitivity of NPV to assumed disease prevalence. Full results appear in **Figure 3**.

## DISCUSSION

Using Bayesian latent class models (BLCM), we estimated a TB prevalence of 14.8% (95% CrI: 11.0–19.3) among children with symptoms in Nairobi, Kenya, significantly higher than the 9.9% identified by culture and molecular testing alone. Our findings align with two published Bayesian Latent Class analyses of pediatric TB. Schumacher et al., analyzing 749 hospitalized South African children with five tests reported moderate culture (60.0%) and geneXpert (49.4%) sensitivity, poor microscopic sensitivity (22.3%), near perfect microbiological specificity, and a sensitive but non-specific CXR (sensitivity 64.2%, specificity 78.0%) (16). Our estimates were similar in direction and magnitude, supporting the robustness of these patterns across the settings. However, our model based prevalence (14.8%) was substantially lower than the (26.7%) reported by Schumacher et al., likely reflecting the differences in case mix, their cohort comprised of exclusively hospitalized children with severe disease, whereas ours included both inpatient and outpatient children and the use of a strongly informative prevalence prior derived from the meta-analysis in our model. Also, our inclusion of the QFT-Plus, which was not available in their panel, further extends the comparison to an immunologic assay not previously evaluated in this framework. A second Latent Class Analysis in a community-based TB screening population (17) similarly reported conditional dependence among microbiological tests, supporting our decision to include random effects in models 2 and 3.

Age was positively associated with TB, consistent with improved diagnostic confirmation in older children. Surprisingly, the point estimate for household TB contact indicated a negative association, though it was not statistically significant as the credible interval crossed the null. Furthermore, the presence of NIH-defined TB signs and symptoms was strongly and significantly negatively associated with the latent TB class (estimate –2.24; 95% CrI: –3.12 to –1.45). This counterintuitive finding potentially reflects the high prevalence of non-specific symptoms (e.g., prolonged cough, fever, weight loss) in the general pediatric population, regardless of underlying etiology, reducing their specificity for TB in this clinical setting.

HIV infection and nutritional indicators (MUAC and WAZ) showed weak and statistically non-significant effects. This contrasts with prior studies linking both HIV and malnutrition to greater TB susceptibility (18) and may be explained by the small numbers of children with HIV and HIV-exposed children in our cohort, both under 10%.

To account for biological dependencies between tests, models 2 and 3 included random effects. A strong, positive dependence was identified among the microbiological tests (culture, geneXpert, smear microscopy), as indicated by a large random effect standard deviation. This suggests test results are highly correlated beyond their shared link to TB status, likely driven by a common underlying factor such as bacillary load. For the immunological tests, a significant non-linear (quadratic) relationship was observed for QFT-Plus , indicating its performance may vary with different levels of disease severity or bacterial burden.

The use of informative priors for microbiological test sensitivities, derived from pediatric meta-analyses, improved precision of posterior estimates for culture and Xpert. By narrowing credible intervals without substantially shifting point estimates, these priors stabilized inference under sparse data conditions. In contrast, vague priors applied to immunological test sensitivities yielded broader credible intervals, whereas specificity estimates remained relatively stable across prior specifications.

Random effects linking microbiological assays and immunological tests improved model fit, acknowledging biological dependencies across diagnostic modalities. Under the assumption of missing at random, Bayesian imputation for missing nutritional status data further strengthened the analysis, ensuring complete case inclusion while propagating uncertainty accordingly.

Key strengths of this study include application of BLCM in a high-burden pediatric setting, incorporation of covariates into prevalence and diagnostic estimates, and handling of missing data using RJAGs-based imputation. These methodological advances allowed for more accurate and complete use of the data compared to conventional analytic approaches.

Our study had limitations. Some covariate estimates, including HIV, were imprecise due to limited numbers in our cohort below 10%. The negative association with household contact, although unexpected, should be interpreted with caution and requires replication. In particular, we cannot rule out the possibility of unmeasured confounding in the estimation of these associations. Finally, while informative priors improved estimation for microbiological tests, results remain sensitive to prior specification.

## Conclusions

This analysis demonstrates the value of BLCMs for generating more accurate estimates of pediatric TB burden and diagnostic performance. A key finding was the estimation of a true TB prevalence of 14.8%, substantially higher than the 9.9% detected by microbiologic confirmation alone, highlighting a significant rate of underdiagnosis in routine care.

The evaluation confirmed that no single diagnostic test is sufficient: culture and Xpert were confirmed as highly specific (>95%) but lacked ideal sensitivity; Chest X-ray demonstrated moderate sensitivity (79.7%) for screening but low specificity; and immunological tests (QFT, TST) showed highly variable and prior-sensitive performance.

Age was identified as a key clinical predictor of TB infection, enhancing test sensitivity in older children, while associations with other covariates like household contact and HIV remained less clear. By successfully integrating prior knowledge, clinical covariates, and random effects to account for dependence, this BLCM framework provides a powerful statistical tool complementing imperfect gold standards for the comprehensive evaluation of TB prevalence and diagnostics in children.

## FUNDING

This work was supported by the NIH/National Institute of Allergy and Infectious Diseases R01AI162152 and R01AI179714, S.M.L., SEATRAC P30-AI168034, TBHTP (Tuberculosis and HIV Co-infection Training Program)- D43-TW011817, J.M.

## AUTHOR CONTRIBUTIONS

S.M.L. designed the parent study. S.M.L.,V.N., G.J.S and E.M.O. developed the parent study protocol. J.M, L.Y.I, S.M.L, G.O.M, performed the data analysis and developed the tables and figures. J.M, L.Y.I, S.M.L, G.O.M, J.N.E, J.K.G, L.A.K, L.N.N prepared the initial draft of the manuscript. All authors have read and approved the manuscript.

## ETHICAL CONSIDERATIONS

Written informed consent was obtained from parents or legal guardians before enrollment. Ethical approvals for the parental study were obtained from the Ethics and Review Committee at the University of Nairobi and Kenyatta National Hospital and the Scientific Ethics and Review Unit at Kenya Medical Research Institute. Secondary analysis of anonymized data did not require additional approval.

## Data Availability

The data underlying this study contain sensitive patient information (including pediatric HIV status) and are subject to ethical restrictions imposed by the University of Nairobi and Kenyatta National Hospital and the Scientific Ethics and Review Unit at Kenya Medical Research Institute. Data cannot be shared publicly. De-identified data are available upon reasonable request to Sylvia M. LaCourse for researchers who meet the criteria for access to confidential data.

## ACKNOWLEDGEMENTS

The authors acknowledge the KEMRI-CRDR study staff, Nairobi County TB clinics, and the referral hospitals. Above all, we offer sincere thanks to the study participants.

## Potential Conflicts of Interest

The authors declare no conflicts of interest.

## Data Access Statement

The data underlying this study contain sensitive patient information (including pediatric HIV status) and are subject to ethical restrictions imposed by the University of Nairobi and Kenyatta National Hospital and the Scientific Ethics and Review Unit at Kenya Medical Research Institute. Data cannot be shared publicly. De-identified data are available upon reasonable request to (Sylvia M LaCourse), for researchers who meet the criteria for access to confidential data.

## Notes

### Competing Interest Statement

The authors have declared no competing interest.

### Author Declarations

University of Nairobi and Kenyatta National Hospital and the Scientific Ethics and Review Unit at Kenya Medical Research Institute

